# Automated detection of genetic relatedness from fundus photographs using Siamese Neural Networks

**DOI:** 10.1101/2023.08.16.23294183

**Authors:** Sakshi Manoj Bhandari, Praveer Singh, Nishanth Arun, Sayuri Sekimitsu, Vineet Raghu, Franziska G. Rauscher, Tobias Elze, Katrin Horn, Toralf Kirsten, Markus Scholz, Ayellet V. Segrè, Janey L. Wiggs, Jayashree Kalpathy-Cramer, Nazlee Zebardast

**Author notes:** **Corresponding Author:** Nazlee Zebardast MD MS, Massachusetts Eye and Ear, Boston, MA. co-first author in alphabetical order (Sakshi Manoj Bhandari, Praveer Singh). co-senior author in alphabetical order (Jayashree Kalpathy-Cramer, Nazlee Zebardast).

## Abstract

Heritability of common eye diseases and ocular traits are relatively high. Here, we develop an automated algorithm to detect genetic relatedness from color fundus photographs (FPs). We estimated the degree of shared ancestry amongst individuals in the UK Biobank using KING software. A convolutional Siamese neural network-based algorithm was trained to output a measure of genetic relatedness using 7224 pairs (3612 related and 3612 unrelated) of FPs. The model achieved high performance for prediction of genetic relatedness; when computed Euclidean distances were used to determine probability of relatedness, the area under the receiver operating characteristic curve (AUROC) for identifying related FPs reached 0.926. We performed external validation of our model using FPs from the LIFE-Adult study and achieved an AUROC of 0.69. An occlusion map indicates that the optic nerve and its surrounding area may be the most predictive of genetic relatedness. We demonstrate that genetic relatedness can be captured from FP features. This approach may be used to uncover novel biomarkers for common ocular diseases.

## Introduction

Heritability, defined as the proportion of overall variation in a population that is attributable to genetic variation, of ocular traits and disease has been extensively studied.^1,2^ Heritability of common eye diseases is relatively high, with glaucoma and age-related macular degeneration (AMD) being two of the most heritable; previous studies have estimated the heritability of glaucoma to be 70% and AMD to be 46-71%.^2,3^ Several fundus features including cup to disc ratio (CDR), disc area, and cup area, have been well examined in family-based studies. CDR are estimated to be 48-79% heritable.^1,4,5^ Disc area and cup area have been found to be 66-77% and 52-83% heritable, respectively.^1,5–7^ Similarly, the high heritability of other ophthalmologic imaging features visualized through optical coherence tomography like ganglion cell complex (GCC) thickness and anterior chamber parameters have been previously documented.^8,9^ The high heritability of ocular traits may indicate a large role of genes or genetic variation in the pathogenesis of ophthalmologic disease.

Color fundus photographs (FPs) capture the retina, optic nerve, and retinal blood vessels which allows for assessment of the interior of the eye. Clear and consistent visualization of nerves and vasculature is unique to FPs and has led to the discovery of novel relationships, including retinal vascular damage and risk of hypertension, as well as diabetic retinopathy and risk of stroke.^10,11^

Prior studies of heritability of ocular traits and diseases have primarily used twins or families. In absence of such information, level of relatedness among a pair of individuals can be calculated from the number of shared alleles, with related individuals sharing more alleles than expected by chance, and the degree of additional sharing proportional to the degree of relatedness.^12^

The recent availability of large-scale genomics data has made it possible to assess relatedness utilizing shared genetics. Large datasets with individuals who may be related but do not necessarily share the same environment can be used to assess the role of heritability and genetics in different imaging features. One example of such a dataset is the UK Biobank (UKBB), a large population-based study composed of greater than 500,000 individuals aged 40-69 across the United Kingdom.^13^ The UKBB contains both genetic and phenotypic data, including fundus photographs, creating a powerful tool to discover new genetic associations of complex traits.

Deep learning is a powerful tool that can be used to automate segmentation and classification of medical images. To date, deep learning models have been used to discover quantitative relationships between fundus appearance and systemic disease, at times succeeding at tasks not previously thought possible, such as predicting age and sex from fundus photographs.^14^ Since fundus photos are more easily acquired than genome data, by harnessing the power of such technology, it may be possible to predict levels of relatedness in subjects where genotype information may not be available and use clustering approaches to uncover novel biomarkers for common heritable eye and systemic diseases.^15,16^

Siamese neural networks, originally designed to verify the authenticity of credit card signatures, take two separate images as inputs and by passing them through twinned neural networks, measure the distance between the two images with respect to their respective imaging features.^17^ Siamese neural networks have been previously used to detect COVID-19 pulmonary disease severity, classify Alzheimer’s disease, assess retinopathy of prematurity from retinal photographs, and assess knee pain from MRI scans, amongst others.^18–21^

In this study, we leverage a Siamese convolutional neural network algorithm for detection of genetic relatedness from FPs using available data from the UKBB and used occlusion maps to identify features of the fundus that are the most inherited. We further use the LIFE-Adult study for external validation of our model. Our approach utilizes the power of big data and deep learning and may offer a way to uncover novel biomarkers in related subjects and sets the stage for better understanding of the pathogenesis of heritable ophthalmologic diseases.

## Materials and methods

### Participants and Datasets

The algorithm was developed using FPs from the UKBB dataset^13^ (http://www.ukbiobank.ac.uk/resources/) and then externally validated on FPs from the LIFE-Adult study.^22^

The UKBB dataset is a prospective cohort study of 502,506 UK residents aged 40-69. Participants were identified from the National Health Service registry. The dataset includes detailed genotypic and phenotypic information on all participants, as well as data from a questionnaire including socio-demographic information, diet, lifestyle, and environmental factors. Over 86,000 people underwent retinal imaging with 45-degree color FPs and paired macular optical coherence tomography (OCT) scans using a Topcon 3D OCT 1000 Mk2 (Topcon, Inc, Japan). The National Research Ethics Service Committee NorthWest–Haydock approved the study, and it was conducted in accordance with the Declaration of Helsinki. All participants provided written informed consent.

The LIFE-Adult study (Leipzig Research Center for Civilization Diseases) is a population-based cohort study of 10,000 randomly selected participants (registry office) from Leipzig, a city with 550,000 inhabitants in the east of Germany. Participants were recruited in an age and sex-stratified manner.^22^ 9,600 participants were in the age group between 40 and 79 years and 400 between 18 and 39 years. This dataset includes detailed genotypic and phenotypic information of participants along with data from a comprehensive examination program that included structured medical interviews, physical and medical examination, questionnaires and cognitive tests including socio-economic status, family medical history, lifestyle, diet, psychological information and sleep.^22^ Over 9,300 people underwent retinal imaging with OCT scans using Heidelberg Spectralis SD-OCT and high quality 45 degree macula centered non-mydriatic fundus imaging using Nidek AFC-230 digital-fundus camera. Of them, 6837 participants were genotyped using the Axiom-CEU microarray. Data were imputed to 1000Genomes phase 3 reference.^23^ Prior to participation, all participants provided written informed consent. The study was approved by the Ethics Committee of the Medical Faculty of the University of Leipzig and was conducted in accordance with the Declaration of Helsinki.

### Genetic relatedness

Array genotype curation process was performed using best practice approaches refined in GTEx consortium. Briefly, participants with unresolved differences between genotype-inferred and reported sex were excluded. Samples with genotyping call rate <97% were removed while variant with call rate < 97% and minor allele frequency (MAF) < 0.01 were removed. We applied Principal Component Analysis (PCA) to linkage disequilibrium (LD)-pruned (r2<0.1 in 200kb windows) genetic markers and the k-nearest neighbors algorithm to predict the ancestral background of participants using ancestral labels from the 1000 Genomes Project Phase 3 reference panel as previously described.^13,24^ The degree of shared ancestry among pairs of UKBB participants was estimated from the kinship coefficient calculated using KING software from a subset of markers weakly aligned to ancestral background, to avoid inflation of the estimate amongst those with mixed ancestry as previously described.^13,24^ This method has been found to have excellent correlation with inference methods implemented in PLINK.^25^ Identity by descent (IBD) was calculated as twice the kinship coefficient. Pairs with IBD > 0.1875 (halfway between the expected IBD for third- and second-degree relatives) but < 0.98 were considered related, while those with non-calculable IBD were considered unrelated.^12^ The upper threshold was chosen to remove duplicates.^13,24^ For the LIFE-Adult dataset, relatedness was calculated in R using an implementation of the estimator for pairwise relatedness of Wang^26^ which is comparable to IBD.

### Image pair labeling

3612 pairs of images (from 2299 different pairs of subjects) from UKBB and 322 pairs of images from LIFE-Adult were labeled as related based on calculated IBD > 0.1875 but < 0.98 as defined above. They were supplemented with an equal number of unrelated pairs, which were generated by taking one of the related pairs’ FP and another one randomly sampled from the rest of the data with a non-calculable IBD for UKBB and IBD < 0.05 for LIFE-Adult. In this work, non-calculable IBD refers to an IBD lower than 0.0884, the lowest calculated IBD in the UKBB cohort. **Figure 1** showcase examples of three FPs from both UKBB and LIFE, (i) and (ii) being related, while (i) and (iii) being unrelated.

**Figure 1.**
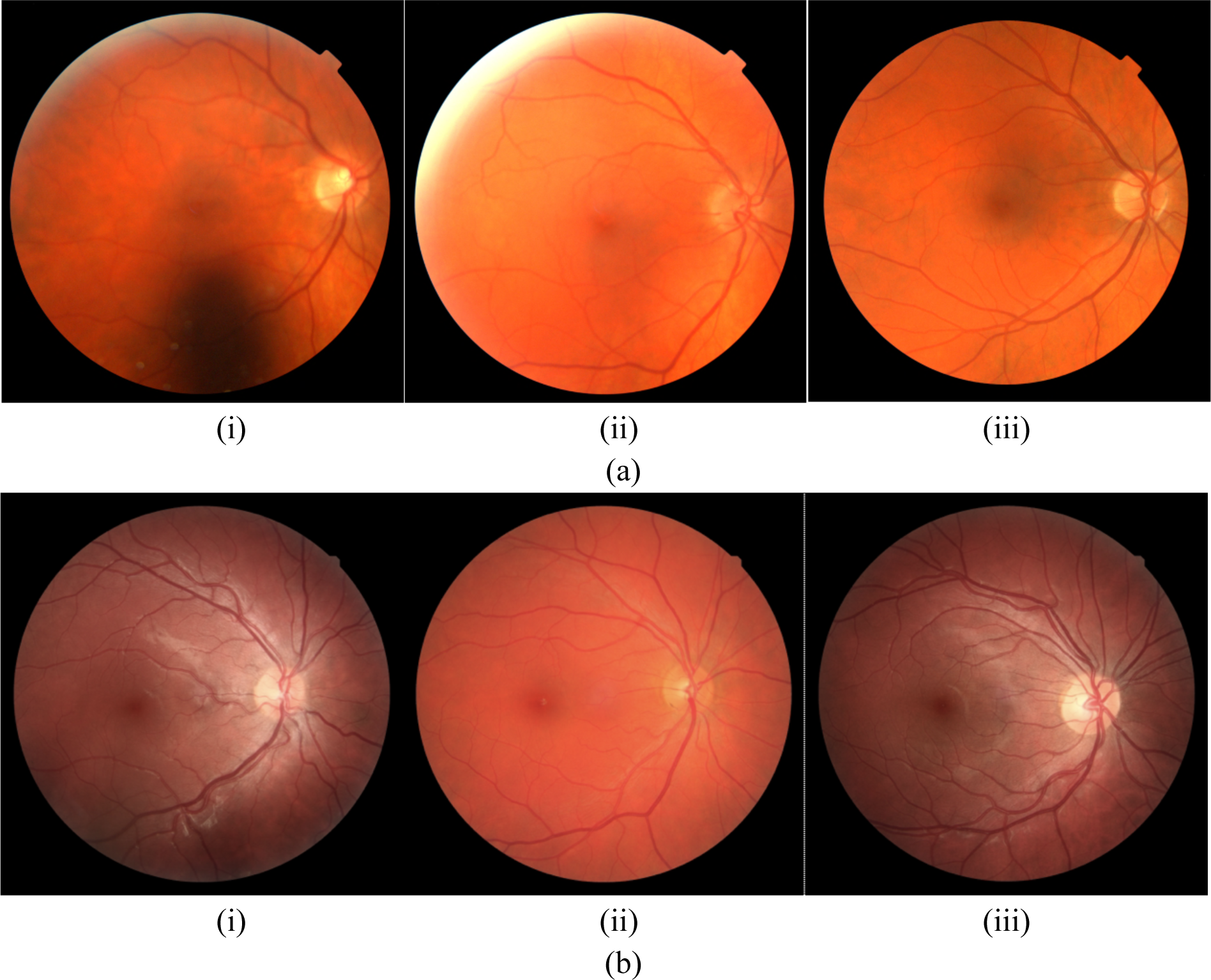
a) Image (i) and (ii) for a related pair from the UKBB dataset and (i) and (iii) is corresponding unrelated pair; b) Image (i) and (ii) for a related pair from the LIFE-Adult dataset and (i) and (iii) is corresponding unrelated pair

### Dataset preparation UKBB dataset

In the UKBB dataset, we had a total of 2299 pairs of related subjects. We created an 80-20 split over this dataset, where 20% related pairs are held out for testing (460 pairs in total) and 80% for training and validation (train-val) (1839 pairs in total). The train-val set was further divided into 10 splits for ten-fold cross validation (each with 183/184 pairs of subjects). All splits were done at subject level, thus ensuring no subject overlap occurs across the different folds. See **Supplemental Figure 1a** for more information. We finally train 10 different models, where for each model, one-fold is used for validation and the other nine folds combined for training. We report the average performance over all 10 models.

Next, to generate pairs for the unrelated cohort, we used all subjects which were not related to any other subject in the UKBB dataset (**Supplemental Figure 1a**). Similar to the related cohort, the unrelated cohort had no subjects overlapping across the 10 different folds. The age difference distribution of the pairs in the related and unrelated cohorts was statistically significant (**Figure 2**). To prevent the model from learning spurious correlations with the age of related and unrelated cohorts, we age-matched the two by sampling the second FP from the rest of the data such that the age difference in the unrelated pair was within ±1 year of the age difference of corresponding related pair (**Supplemental Figure 1b**). **Figure 2** shows the age distribution of the age matched and non-age matched cohorts for both UKBB as well as LIFE-Adult dataset.

**Figure 2.**
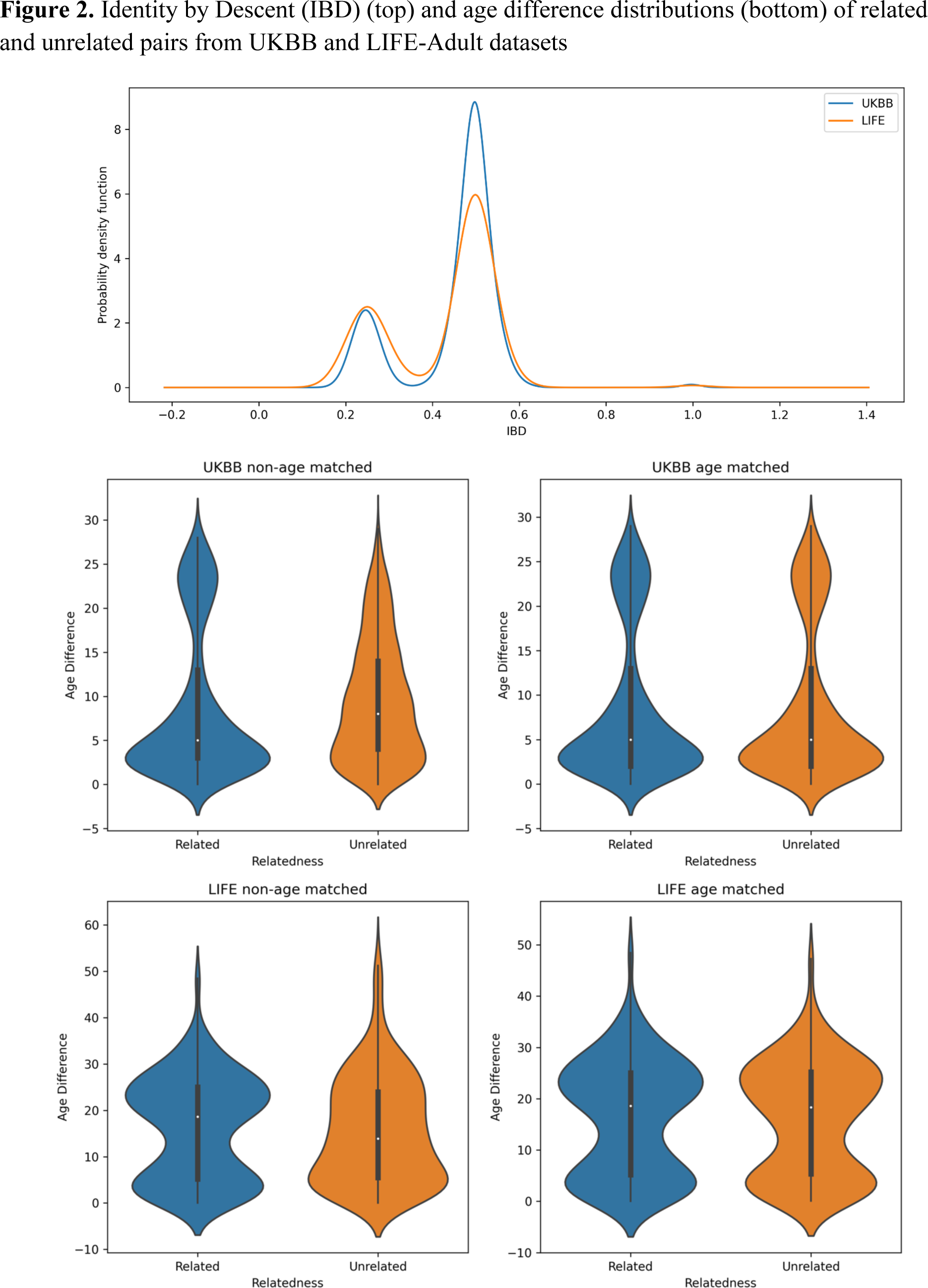
Identity by Descent (IBD) (top) and age difference distributions (bottom) of related and unrelated pairs from UKBB and LIFE-Adult datasets

### LIFE-Adult dataset

We identified 448 subjects (224 pairs of subjects from 218 families) out of 7669 participants in LIFE-Adult who were related with IBD > 0.1875. Similar to the UKBB dataset, we randomly selected unrelated controls (using IBD < 0.05) by sampling from over 6000 participants, matched by sex and age relative to the cases.

The complete LIFE-Adult dataset was first used as an external test set on models trained entirely on UKBB dataset. Next, the LIFE-Adult dataset was divided into 10 splits (each split consisting of 64/66 pairs of subjects). These splits were subsequently used in the 10-fold cross validation when we transfer learn on LIFE-Adult, using the best performing model trained on UKBB. Transfer learning is a commonly used technique in machine learning where knowledge gained from one task is utilized for a different, but related task. In this case, we used the model weights learnt after training on the UKBB dataset to fine-tune over the LIFE-Adult dataset.

### Siamese Convolutional Network and Classifier

Convolutional Neural Networks (CNNs) are artificial intelligence models largely used for image classification, object detection and segmentation.^27^ Siamese Neural Networks are a special class of CNNs wherein we can compute similarity between two input images by comparing feature vector outputs from two identical branches of the network that share parameters. In this analysis, we used DenseNet121 networks as backbones for the identical branches in our Siamese network (see **Figure 3** for an overview of our approach).^28^ Densenet121 is a CNN architecture that consists of four blocks with 6, 12, 24 and 16 layers each, where each later block has access to information from all the previous blocks through dense connections, thus ensuring maximum information flows to the penultimate layer and ultimately resulting in a robust feature extractor. The output feature vectors (q and p) corresponding to the two input images are extracted from each DenseNet121 branch and compared using Euclidean Distance (ED). A smaller value of ED (compared to a set threshold) implies pairs are related, and unrelated otherwise.

**Figure 3.**
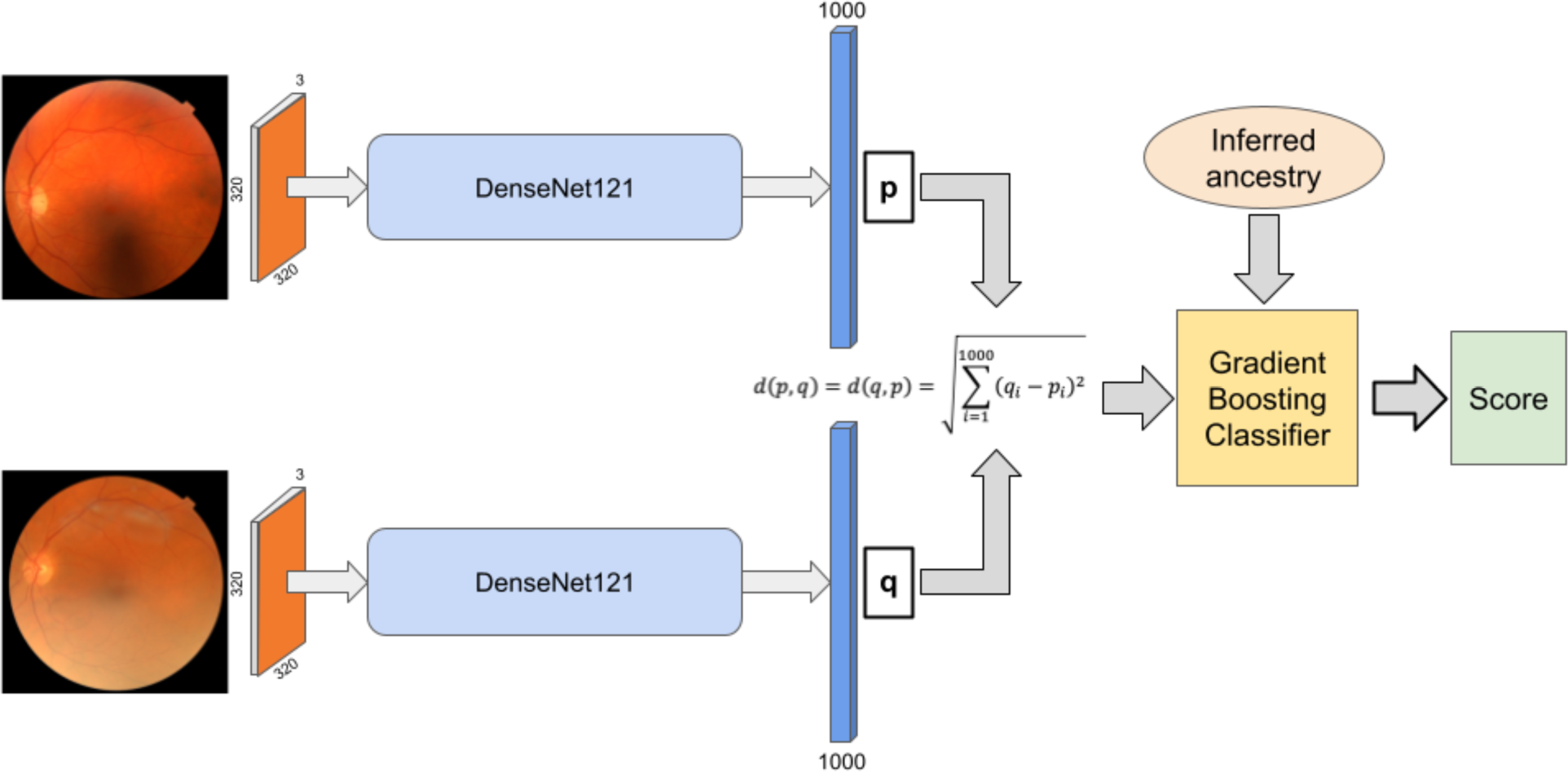
Overview of the proposed algorithm. Pair of retinal fundus photographs (RFPs) are fed through the Siamese Network to output a Euclidean distance b/w the two RFPs. This distance combined with the inferred ancestry label is then fed to a gradient-booster classifier to finally output a relatedness score.

The Siamese network is trained with the Adam optimization algorithm and contrastive loss, which is designed to separate unrelated pairs of images and unite related pairs of images in output feature space.^29,30^ The contrastive loss function is given by:

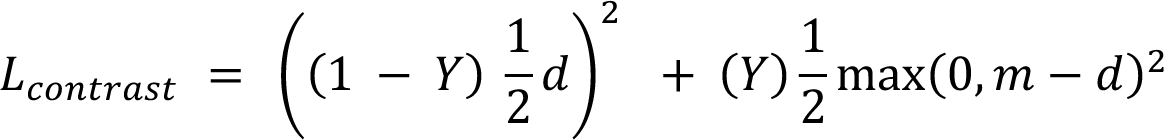

where Y is the relatedness label, d is the computed Euclidean Distance and m is a hyperparameter used to control the extent of separation between classes.

As a result of this contrastive loss, related pairs of images are expected to produce a smaller Euclidean Distance between the output arms of the Siamese network as compared to unrelated pairs. An optimal threshold for Euclidean Distance can then be computed using Youden Index to differentiate between related and unrelated pairs during inference.^31^ The model was trained with a learning rate of 0.0001 and early stopping to avoid overfitting with a patience of 4 epochs while monitoring the validation loss.^32^ To standardize right and left eyes, all left eye images underwent a horizontal flip as a preprocessing step, which were then resized to a standard size of 320 x 320.

The raw Euclidean Distance obtained with the Siamese network was further combined with an indicator for inferred ancestry in the two fundus photographs. This integration was achieved using a Gradient Boosting Classifier, which combined the Euclidean Distance with inferred ancestry to produce a final probability of genetic relatedness.^33^ This classifier was trained with 100 boosting estimators and a learning rate of 0.1.

### *10-fold* Cross-Validation

For the UKBB dataset, we performed a 10-fold cross validation, where nine folds were used for training and one for validation. We finally compute average AUROC along with 95% confidence interval (CI) over the 10-folds. Final testing on the held-out UKBB test set as well as on the full external LIFE-Adult dataset was done using model ensembling, wherein we averaged the normalized Euclidean Distances obtained from the 10 models and then calculated the AUROC.

Additionally, we also experimented by fine-tuning the best model (closest in performance to model ensembling) from UKBB over the LIFE-Adult dataset after reducing the learning rate by 1/10 and updating all model weights during the fine-tuning process. In order to conduct this fine-tuning experiment on LIFE-Adult, we divided LIFE-Adult into 10 folds (similar to UKBB 10-fold cross validation) and reported the average AUROC and 95% CI for the LIFE-Adult dataset.

### Model Interpretability

For better visualization of the underlying factors influencing predictions from the Siamese model, we employ an occlusion map technique, wherein different regions of the input image are selectively occluded to observe the corresponding change in the output.^34^ This produces a heatmap, where regions of the image are highlighted in proportion to the extent of change in output ED by occluding those regions, thereby offering post-hoc interpretability of the AI model. Although occlusion maps are well defined and used in the single image classification setting, we extend it to our pairwise learning task by simultaneously occluding corresponding patches of size 32 x 32 on both FPs to observe areas that lead to the highest magnitude of change in the computed Euclidean Distance.

To understand the relative contribution of fundus vasculature pattern on model prediction, we used a UNet trained on the DRIVE, ChaseDB and STARE retinal fundus photograph databases on the training, validation, and test splits (https://lmb.informatik.uni-freiburg.de/people/ronneber/u-net/).^35^ The resulting segmentations (**Supplemental Figure 2a**) were used as inputs to the Siamese network in place of the fundus photographs in the classification task.

### Statistical analyses & Software Tools

Statistical analyses were performed using NumPy and Pandas. The Euclidean Distance between the outputs of the arms of the Siamese network was taken as the final prediction to compute the Area under the Receiver Operator Curve (AUROC) curve of the model with a binary indicator of relatedness as the ground truth label. The model was evaluated by calculating sensitivity and specificity at different ED thresholds and subsequently plotting the AUROC curve, all of which were calculated using the inbuilt scikit-learn metrics. For inference, an optimal threshold was chosen via Youden Index to differentiate between related and unrelated pairs. All the pairs with ED less than the cut-off for the optimal Youden Index were classified as related and the rest as unrelated. Average AUROC and 95% confidence interval reported over the 10-folds of UKBB and LIFE-Adult validation sets were computed using the scipy stats package.

We also plotted a Uniform Manifold Approximation and Projection (UMAP) for UKBB versus LIFE-Adult dataset (**Figure 4**) using Python UMAP library, where the image features were reduced to 2 dimensions. UMAP is a dimension reduction technique that can be used for high-dimensional model feature visualization or other general non-linear dimension reduction.

**Figure 4.**
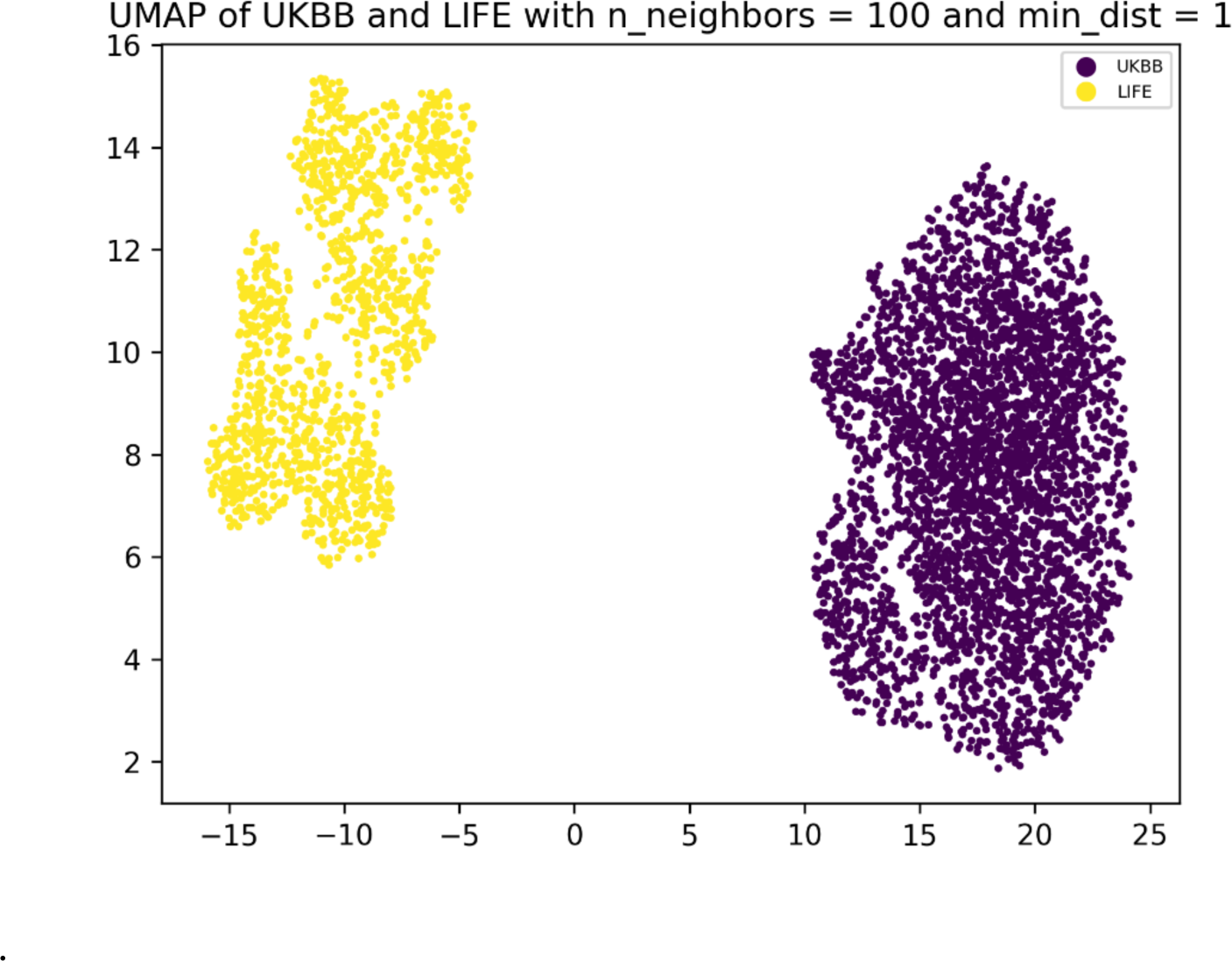
UMAP of UKBB versus LIFE-Adult images demonstrating two different populations. UMAP provides a lower dimension representation of the samples for better visualization.

## Results

**Table 1** summarizes age-matched cohort characteristics for train+val and test sets of UKBB as well as the LIFE-Adult dataset. The distribution of age for both non-age matched and age-match cohorts of UKBB and LIFE-Adult datasets and the distribution of IBD scores for the related cohort for the two datasets is shown in **Figure 2**.

**Table 1.** Training Cohort characteristics for both UKBB and Life-Adult Datasets.

| Attribute | Age matched (Train + Val) |  | Age matched (Test) |  |
| --- | --- | --- | --- | --- |
|  | Related Cohort | Unrelated cohort | Related Cohort | Unrelated cohort |
| UKBB Dataset |  |  |  |  |
| Total number of pairs (pairs/participants) | 3612/7224 | 3612/7224 | 903/1806 | 903/1806 |
| Image pairs from same gender (%/pairs) | 57.2/2066 | 50.2/1814 | 56.1/507 | 45.5/411 |
| Female participants (%/participants) | 59.2/4276 | 54.9/3964 | 56.6/1022 | 55.0/994 |
| Image pairs from same ancestry (predicted) (%/pairs) | 99.8/3606 | 83.8/3028 | 100/903 | 85.3/716 |
| Age (years), mean (SD) | 56.6 (8.7) | 56.6 (8.9) | 56.7 (8.6) | 56.5 (8.7) |
| Age difference for image pairs (years), mean (SD) | 9.8 (8.5) | 9.8 (8.4) | 9.3 (8.1) | 9.3 (8.2) |
| LIFE-Adult Dataset |  |  |  |  |
| Total number of pairs (pairs/participants) | 322/644 | 322/644 |  |  |
| Image pairs from same gender (%/pairs) | 49.37/159 | 49.37/159 |  |  |
| Female participants (%/participants) | 57.0/367 | 57.0/367 |  |  |
| Age (years), mean (SD) | 55.7 (13.7) | 55.7 (13.7) |  |  |
| Age difference for image pairs (years), mean (SD) | 16.3 (11.0) | 16.3 (10.9) |  |  |

### Model Performance

For the age-matched UKBB dataset, the Siamese network yields a mean AUROC of 0.9103 (95% CI = 0.9028,0.9177) on the 10-fold cross validation and 0.9262 on the held-out UKBB test dataset using the ensemble model (**Figure 5a**). **Figure 5b** shows a violin plot illustrating the separation of the two classes in output feature space for the model trained on the age matched dataset. The mean and standard deviation of the ED predictions for the related class on the test set were 19.09 and 11.86, while corresponding values for the unrelated class were 40.17 and 14.46 (**Supplementary Table 1**). **Figure 6b** shows two examples of incorrect classifications by the model.

**Figure 5.**
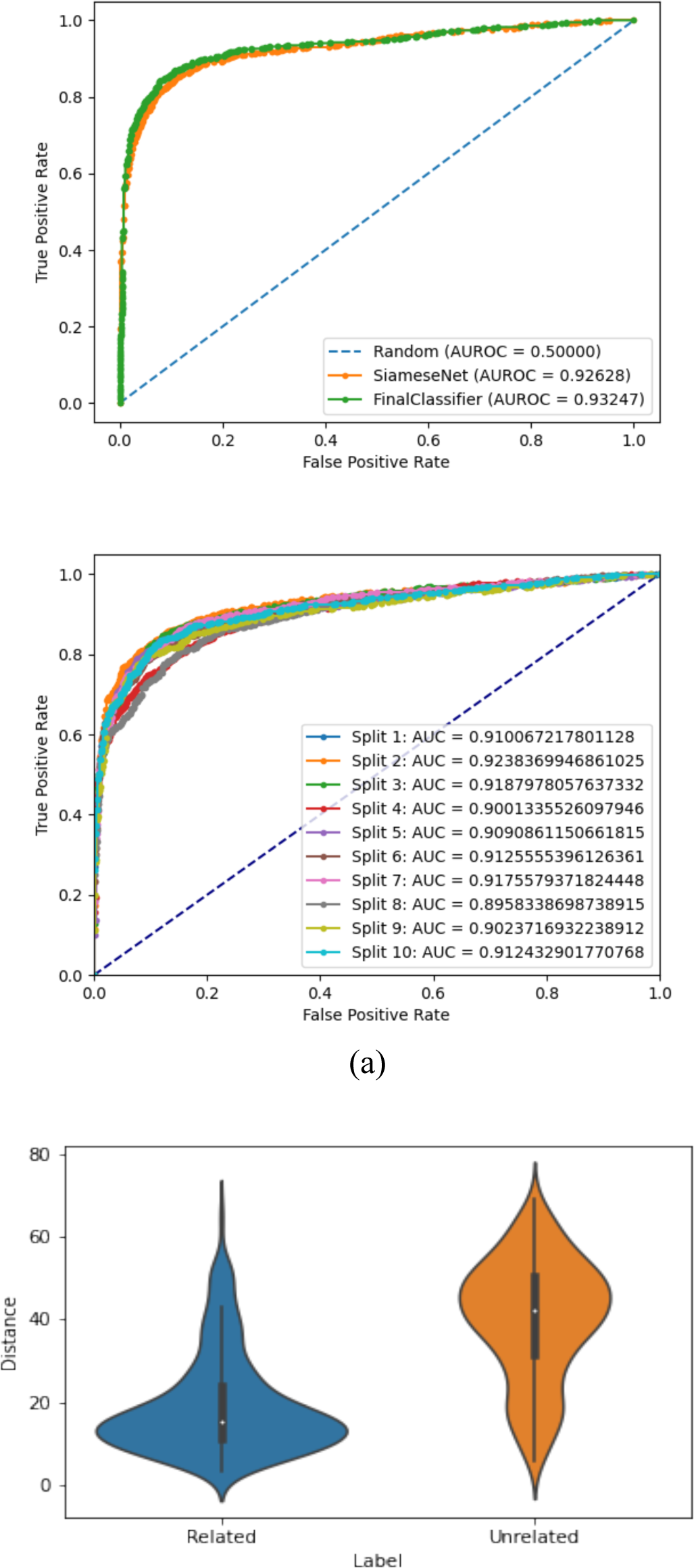
(a) ROC Curve for (i) the Siamese network and the final classifier combining Euclidean Distance and inferred ancestry and (ii) the tenfold cross validation on UKBB dataset; (b) Violin plot showing the distribution of Euclidean Distance scores for both classes in UKBB dataset.

**Figure 6.**
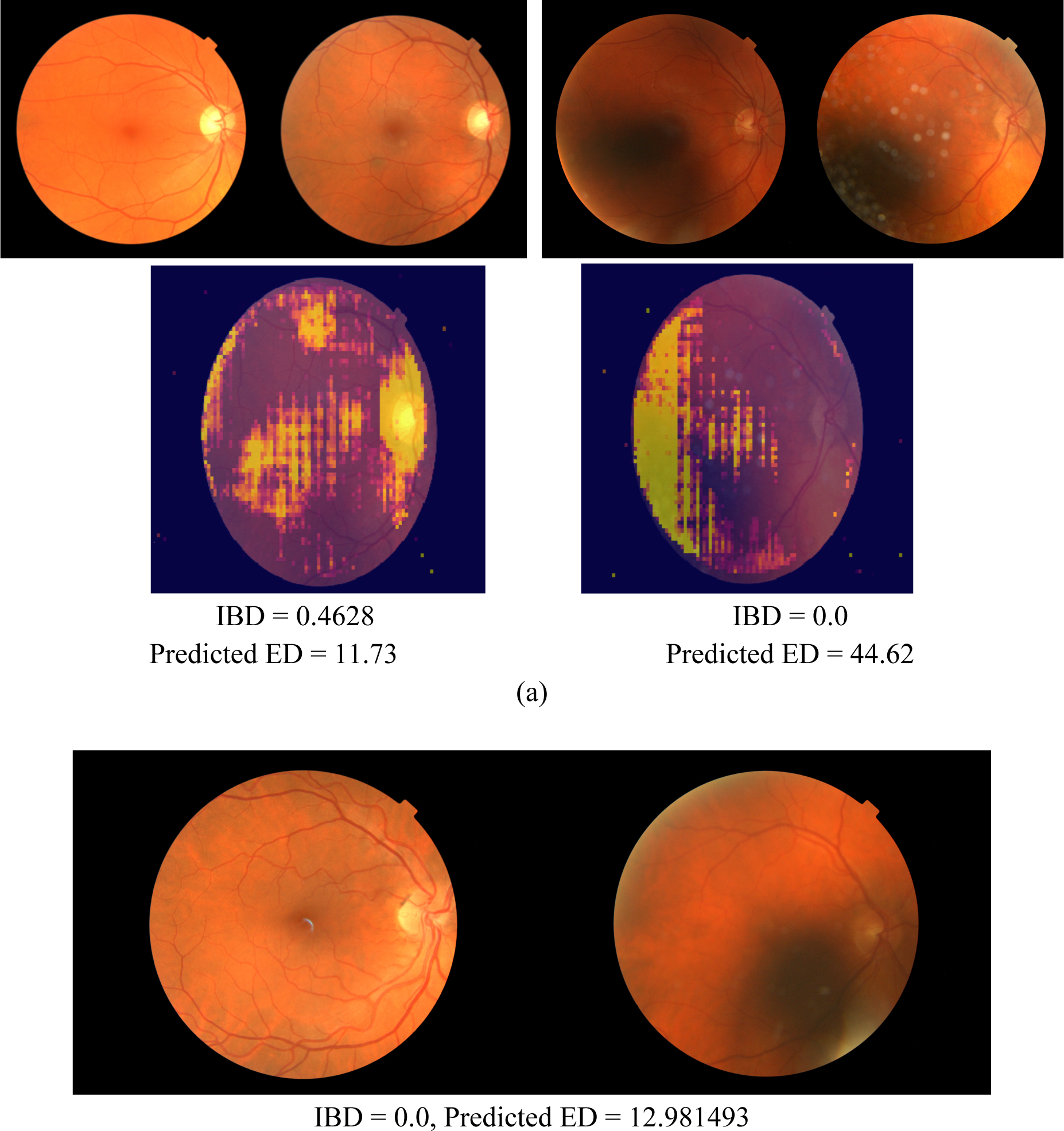
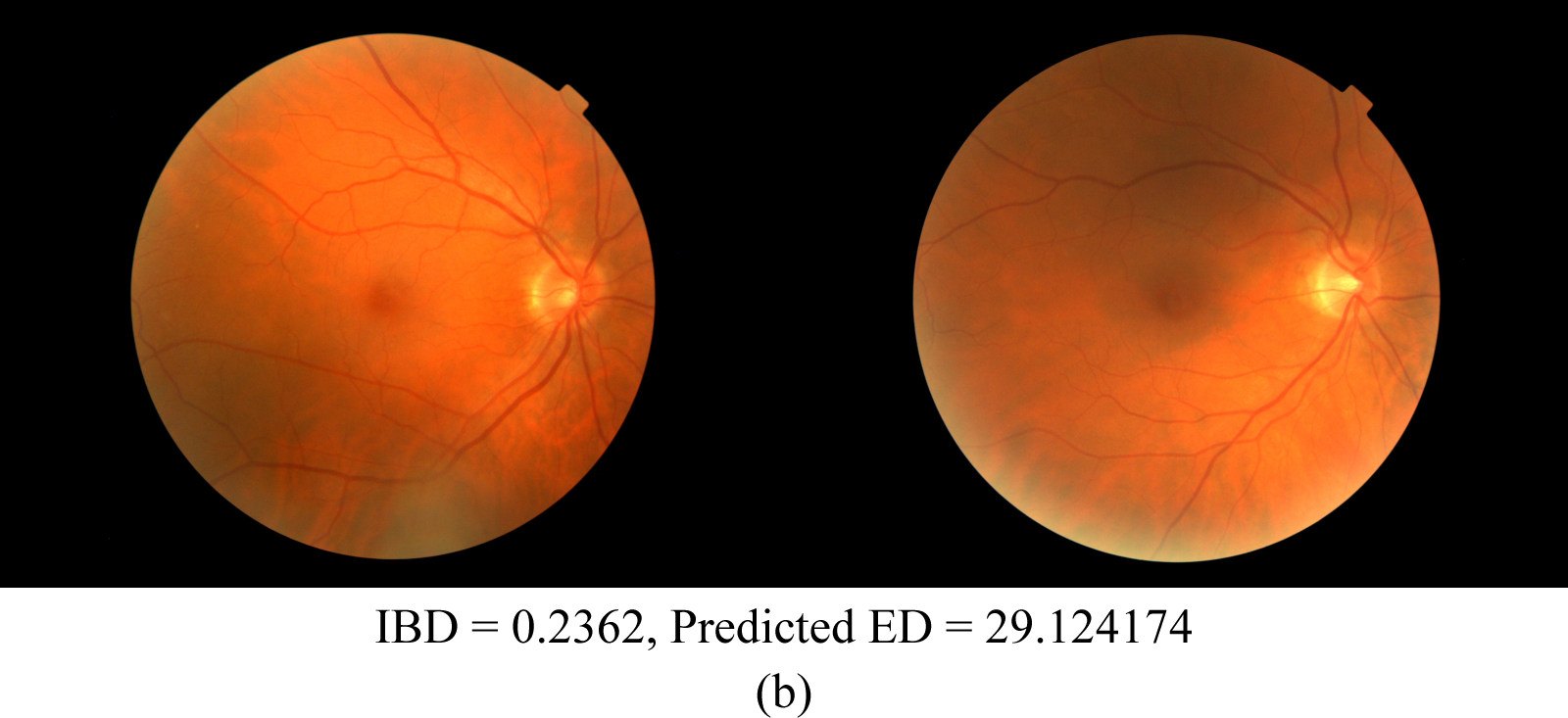
(a) Occlusion maps generated on a pair of related images (left) and unrelated images (right) where yellow regions indicate higher importance for prediction (Threshold Euclidean Distance = 17.895151; lower predicted Euclidean Distance = higher relatedness while lower IBD = lower relatedness) (b) Misclassified Examples: Example of an unrelated pair classified incorrectly as related (top) and a related pair classified incorrectly as unrelated (bottom) for the UKBB dataset (Threshold Euclidean Distance = 17.895151; lower ED = higher relatedness while lower IBD = lower relatedness)

For the UKBB dataset, model corresponding to fold-2 was determined to be the best performing model from the tenfold cross validation (**Supplementary Table 1**), since it was closest to the ensemble model performance on the held out UKBB test set, with an AUROC of 0.9238. These ten models trained on UKBB when tested on the complete LIFE-Adult dataset resulted in an AUROC of 0.5815, using the ensemble model. The best forming model from fold-2 UKBB dataset alone resulted in an AUROC of 0.5545. The same model when fine-tuned over the LIFE-Adult dataset, through a 10-fold cross-validation (like UKBB dataset) resulted in a mean AUROC of 0.5935 (95% CI = 0.549, 0.638) (**Figure 1, Supplementary Table 2**).

To determine the potential cause for poorer performance on the LIFE-Adult dataset, we plotted a UMAP for UKBB versus LIFE-Adult (**Figure 4**), where the image features were reduced to 2 dimensions. The UMAP demonstrated two distinct image clusters corresponding to UKBB and LIFE-Adult datasets, highlighting wide differences in the distributions of the two datasets. We tried improving model performance on LIFE-Adult dataset by applying local color normalization as well as color jitters to the images before training to discount the effects of the differences in the saturation and hue between UKBB and LIFE-Adult images (**Figure 1**), but the results did not improve.

### Model Interpretability using Occlusion Maps and Vessel Segmentation

**Figure 6a** illustrates an occlusion map generated from the best performing age matched UKBB model. The occlusion map showcases that the optic nerve, and the surrounding area may be the most predictive of the model decision. Of note, in the related pairs of images, the occlusion of space corresponding to the optic nerve seemed to play a critical role in the output of the Siamese network whereas for unrelated pairs, the obtained heatmap is more diffused, indicating that a larger attention is placed on regions outside the optic nerve area.

In order to test how relevant the vascular structures independently are to predict generated relatedness, we trained the Siamese network on vessel segmentations in place of raw fundus photographs, resulting in an AUROC of 0.75 (**Supplementary** Figure 2). When the same model was tested on the LIFE-Adult study, the AUROC was 0.526. This drop-off suggests that that the optic nerve rather than major fundus vessel patterns may be a more salient region in the fundus photographs for the neural network’s genetic relatedness classification performance.

Alternatively, the finer branching details of the vasculature, which were often not accurately shown in the segmentations, may play an important role in determining genetic relatedness. This is further supported by the occlusion maps which also highlighted regions outside the optic nerve.

### Final Classification

When an indicator for inferred ancestry was incorporated into the prediction by combining it with the obtained Euclidean Distance using a gradient boosting classifier, the AUROC metric was 0.9324, demonstrating slight improvement over the raw Siamese network prediction. Figure 5a shows the ROC curves for the combined classifier and the Siamese network respectively trained on the age matched UKBB dataset.

## Discussion

We found that Euclidean Distances derived from a Siamese neural network were able to accurately determine genetic relatedness from two fundus photographs, with an AUROC of 0.93 (addition of inferred ancestry showing marginal improvement in performance). The occlusion map produced by the model indicates that the optic nerve is highly predictive of the model’s decision. The UNet blood vessel segmentation did not improve prediction performance of the model. This may indicate that both the optic nerve as well as the finer branching of retinal vasculature, not always present in segmentation, may play an important role in determining genetic relatedness.

Our findings indicate high heritability of fundus features as well as conservation of optic nerve features among relatives. Prior work has demonstrated the high heritability of ocular structures, particularly the optic disc. Healey et al. previously found that heritability of the optic disc area was 0.73 and the optic cup area was 0.66.^6^ Han et al. similarly demonstrated high heritability of vertical cup-to-disc ratio, the presence of peripapillary atrophy to be highly heritable; this result was consistent with previous work.^1,36^ Indeed, to date, in GWAS studies several genes and single-nucleotide polymorphisms (SNPs) have been associated with optic disc area or vertical cup-to-disc ratio.^37,38^ The nearest genes to the associated SNPs include CDKN2b-AS1, SIX6, BMP2, FLNB, amongst others. To this end, we found that in related pairs, the region surrounding the optic nerve seemed to play a major role in the output of the Siamese network, while in unrelated pairs, the heatmap is more diffused. We plan to extend our occlusion and saliency analysis in future experiments to further understand what might be causing this observation. Future exploration in this area may shed light on novel pathways of glaucoma pathogenesis.

While our model trained on vessel segmentations performed less well than the model trained on the entire fundus photograph, blood vessel patterns demonstrated reasonable concordance among related pairs with AUROC of 0.75. Retinal vascular features have been previously found to be highly associated with systemic diseases or their risk factors, like hypertension and coronary heart disease.^39–41^ These features include arteriole narrowing, branching patterns, and vessel tortuosity. Kirin et al. found several of these features to be heritable; arterial tortuosity and venular tortuosity were found to have a heritability of 0.55 and 0.21 respectively.^42^ Similarly, Liu et al. found heritability of central retinal arteriole equivalent and central retinal venule equivalent to be 0.21 and 0.34, indicating moderate heritability.^43^ While it is possible that vascular features may be less conserved among related individuals relative to the optic nerve features, the drop in accuracy of our model may also indicate that the smaller branching of retinal vessels, those not captured by our segmentation, may play an important role in determining genetic relatedness.

There are several age-related diseases that have a vascular component and may impact the appearance of retinal vessels. Lemmens et al. previously found that retinal vasculature complexity increases with age.^44^ To assure that age differences were not driving the results we found, we tested our model using both an age matched and non-age matched unrelated cohort. We found that performance improved slightly on the test set after age matching the related and unrelated cohorts. Similarly, we found that adding inferred ancestry information to the ED computed from fundus photograph features also slightly improved the results. Previous research has found differences in optic disc size amongst different ethnic groups. However, the majority of individuals included in this study were of European descent, likely minimizing the impact of ancestry on the performance of the model.^45^ Overall, these findings indicate that while age and inferred ancestry can contribute to retinal features, there are other factors that are driving our model’s predictive accuracy.

Our model demonstrated good internal validity with AUROC ranging from 0.89 to 0.924 on our 10-fold cross validation experiments. However, The AUROC values on the LIFE-Adult dataset were lower than expected. We attribute this to two reasons. First, the optic disk in LIFE-Adult images is slightly to the left as compared to the UKBB images (Figure 1). Since all the features surrounding the optic disk and vessels were important, we did not do any random cropping or affine transformation while training and hence the model was not robust to the positioning of the optic disk. Second, the colorations and saturation of the LIFE-Adult images were quite different from the UKBB (Figure 1), which may have affected the performance of our model. More importantly, we noted that the low AUROC on LIFE-Adult was largely because the model was predicting most of the samples as related (reflected in **Supplemental Table 2** where for each Life-Adult fold, the calibrated average ED of unrelated samples is small and close enough to the average ED of related samples). There is not only low genetic differentiation among Europeans, but there is a close correspondence between genes and geographies in Europe.^46^ Since all the participants of the LIFE-Adult study were from the same town in Germany, the genetic differentiation among the participants might not have been sufficient for the model. This effect could be compounded by the considerable sample size limitation in LIFE-Adult following fine-tuning.

Our study is subject to several limitations. First, due to the limited number of related individuals in our dataset, we combined everyone with IBD >0.1875 into a binary variable of related vs unrelated pairs. In the future, use of a larger dataset may allow us to use a similar model to predict levels of genetic relatedness, allowing for better understanding of model decision making. Another limitation of our work was the inability to segment smaller, finer vessels. As a result, we may be underestimating the importance of blood vessel patterns in predicting genetic relatedness. Finally, the UKBB and LIFE-Adult are both composed of predominantly healthy individuals of primarily European ancestry and is not fully representative of other populations. Future work in more diverse datasets will be critical to confirm the reproducibility of this work in other populations.

In conclusion, we demonstrate that a Siamese neural network can be used to accurately determine genetic relatedness from fundus photographs. Our novel findings suggest that deep learning can be used as an important tool to detect conserved genetic ocular features in different disease states. Future exploration in larger datasets may uncover critical novel biomarkers to help us better understand pathogenesis of ocular disease in related patients.

## Data Availability

Please contact authors for availability of data produced in present study.

## Meeting presentation

This work was presented at Association for Research in Vision and Ophthalmology (ARVO) 2021, virtual conference.

## References

1. Sanfilippo, P. G., Hewitt, A. W., Hammond, C. J. & Mackey, D. A. The Heritability of Ocular Traits. Survey of Ophthalmology 55, 561–583 (2010).

2. Wang, K., Gaitsch, H., Poon, H., Cox, N. J. & Rzhetsky, A. Classification of common human diseases derived from shared genetic and environmental determinants. Nat Genet 49, 1319–1325 (2017).

3. Seddon, J. M. The US Twin Study of Age-Related Macular Degeneration: Relative Roles of Genetic and Environmental Influences. Arch Ophthalmol 123, 321 (2005).

4. Klein, A. P. Heritability Analysis of Spherical Equivalent, Axial Length, Corneal Curvature, and Anterior Chamber Depth in the Beaver Dam Eye Study. Arch Ophthalmol 127, 649 (2009).

5. He, M. et al. Heritability of optic disc and cup measured by the Heidelberg Retinal Tomography in Chinese: the Guangzhou twin eye study. Invest Ophthalmol Vis Sci 49, 1350–1355 (2008).

6. Healey, P. et al. The heritability of optic disc parameters: a classic twin study. Invest Ophthalmol Vis Sci 49, 77–80 (2008).

7. van Koolwijk, L. M. E. et al. Genetic Contributions to Glaucoma: Heritability of Intraocular Pressure, Retinal Nerve Fiber Layer Thickness, and Optic Disc Morphology. Invest. Ophthalmol. Vis. Sci. 48, 3669 (2007).

8. Bloch, E. et al. Genetic and Environmental Factors Associated With the Ganglion Cell Complex in a Healthy Aging British Cohort. JAMA Ophthalmol 135, 31 (2017).

9. Ciociola, E. C. et al. The Heritability of Primary Angle Closure Anatomic Traits and Predictors of Angle Closure in South Indian Siblings. American Journal of Ophthalmology S0002939421002804 (2021) doi:10.1016/j.ajo.2021.04.038.

10. Drinkwater, J. J. et al. Retinopathy predicts stroke but not myocardial infarction in type 2 diabetes: the Fremantle Diabetes Study Phase II. Cardiovasc Diabetol 19, 43 (2020).

11. Huang, K. et al. The association between retinal vessel abnormalities and H-type hypertension. BMC Neurol 21, 6 (2021).

12. Anderson, C. A. et al. Data quality control in genetic case-control association studies. Nat Protoc 5, 1564–1573 (2010).

13. Bycroft, C. et al. The UK Biobank resource with deep phenotyping and genomic data. Nature 562, 203–209 (2018).

14. Korot, E. et al. Predicting sex from retinal fundus photographs using automated deep learning. Sci Rep 11, 10286 (2021).

15. Slieker, R. C. et al. Distinct Molecular Signatures of Clinical Clusters in People With Type 2 Diabetes: An IMI-RHAPSODY Study. Diabetes 70, 2683–2693 (2021).

16. Isik, Z. et al. In silico identification of novel biomarkers for key players in transition from normal colon tissue to adenomatous polyps. PLoS ONE 17, e0267973 (2022).

17. Bromley, J. et al. SIGNATURE VERIFICATION USING A “SIAMESE” TIME DELAY NEURAL NETWORK. Int. J. Patt. Recogn. Artif. Intell. 07, 669–688 (1993).

18. Li, M. D. et al. Automated assessment of COVID-19 pulmonary disease severity on chest radiographs using convolutional Siamese neural networks. medRxiv 2020.05.20.20108159 (2020) doi:10.1101/2020.05.20.20108159.

19. Mehmood, A., Maqsood, M., Bashir, M. & Shuyuan, Y. A Deep Siamese Convolution Neural Network for Multi-Class Classification of Alzheimer Disease. Brain Sci 10, E84 (2020).

20. Chang, G. H. et al. Assessment of knee pain from MR imaging using a convolutional Siamese network. Eur Radiol 30, 3538–3548 (2020).

21. Li, M. D. et al. Siamese neural networks for continuous disease severity evaluation and change detection in medical imaging. npj Digit. Med. 3, 48 (2020).

22. Loeffler, M. et al. The LIFE-Adult-Study: objectives and design of a population-based cohort study with 10,000 deeply phenotyped adults in Germany. BMC Public Health 15, 691 (2015).

23. Pott, J. et al. Genome-wide meta-analysis identifies novel loci of plaque burden in carotid artery. Atherosclerosis 259, 32–40 (2017).

24. Manichaikul, A. et al. Robust relationship inference in genome-wide association studies. Bioinformatics 26, 2867–2873 (2010).

25. Purcell, S. et al. PLINK: a tool set for whole-genome association and population-based linkage analyses. Am J Hum Genet 81, 559–575 (2007).

26. Wang, J. An estimator for pairwise relatedness using molecular markers. Genetics 160, 1203–1215 (2002).

27. Krizhevsky, A., Sutskever, I. & Hinton, G. E. ImageNet classification with deep convolutional neural networks. Commun. ACM 60, 84–90 (2017).

28. Huang, G., Liu, Z., van der Maaten, L. & Weinberger, K. Q. Densely Connected Convolutional Networks. *arXiv:1608.06993 [cs]* (2018).

29. Chopra, S., Hadsell, R. & LeCun, Y. Learning a Similarity Metric Discriminatively, with Application to Face Verification. in 2005 IEEE Computer Society Conference on Computer Vision and Pattern Recognition (CVPR’05) vol. 1 539–546 (IEEE, 2005).

30. Kingma, D. P. & Ba, J. Adam: A Method for Stochastic Optimization. *arXiv:1412.6980 [cs]* (2017).

31. Ruopp, M. D., Perkins, N. J., Whitcomb, B. W. & Schisterman, E. F. Youden Index and optimal cut-point estimated from observations affected by a lower limit of detection. Biom J 50, 419–430 (2008).

32. Caruana, R., Lawrence, S. & Giles, L. Overfitting in Neural Nets: Backpropagation, Conjugate Gradient, and Early Stopping. in vol. 13 381–387 (MIT Press, 2000).

33. Ke, G., Qi, M., Finley, T., Wang, T. & Chen, W. LightGBM: A Highly Efficient Gradient Boosting Decision Tree. in (2017).

34. Hou, Y.-L., Peng, J., Hao, X., Shen, Y. & Qian, M. Occlusion localization based on convolutional neural networks. in 2017 IEEE International Conference on Signal Processing, Communications and Computing (ICSPCC) 1–5 (IEEE, 2017). doi:10.1109/ICSPCC.2017.8242508.

35. Ronneberger, O., Fischer, P. & Brox, T. U-Net: Convolutional Networks for Biomedical Image Segmentation. arXiv:1505.04597 [cs] (2015).

36. Han, J. C. et al. Heritability of the morphology of optic nerve head and surrounding structures: The Healthy Twin Study. PLoS One 12, e0187498 (2017).

37. Ramdas, W. D. et al. A genome-wide association study of optic disc parameters. PLoS Genet 6, e1000978 (2010).

38. Springelkamp, H. et al. New insights into the genetics of primary open-angle glaucoma based on meta-analyses of intraocular pressure and optic disc characteristics. Hum. Mol. Genet. ddw399 (2017) doi:10.1093/hmg/ddw399.

39. Liew, G., Wang, J. J., Mitchell, P. & Wong, T. Y. Retinal vascular imaging: a new tool in microvascular disease research. Circ Cardiovasc Imaging 1, 156–161 (2008).

40. MacGillivray, T. J. et al. Suitability of UK Biobank Retinal Images for Automatic Analysis of Morphometric Properties of the Vasculature. PLoS One 10, e0127914 (2015).

41. McGowan, A. et al. Evaluation of the Retinal Vasculature in Hypertension and Chronic Kidney Disease in an Elderly Population of Irish Nuns. PLoS One 10, e0136434 (2015).

42. Kirin, M. et al. Determinants of retinal microvascular features and their relationships in two European populations. J Hypertens 35, 1646–1659 (2017).

43. Liu, Y.-P. et al. Heritability of the retinal microcirculation in Flemish families. Am J Hypertens 26, 392–399 (2013).

44. Lemmens, S. et al. Age-related changes in the fractal dimension of the retinal microvasculature, effects of cardiovascular risk factors and smoking behaviour. Acta Ophthalmol (2021) doi:10.1111/aos.15047.

45. Lee, R. Y. et al. Ethnic variation in optic disc size by fundus photography. Curr Eye Res 38, 1142–1147 (2013).

46. Novembre, J. et al. Genes mirror geography within Europe. Nature 456, 98–101 (2008).

