## Supplement for "Automated detection of genetic relatedness from fundus photographs using Siamese Neural Networks"

**Supplementary material:**

**Supplementary Table 1.** Table showing for the 10 different cross-validation folds, corresponding train and val losses, Avg Euclidean Distances of Related vs Unrelated cohorts as well as AUROC's on the Validation and held out test set of UKBB dataset.

| Fold | TRAIN | VAL | | | | TEST |
|---|---|---|---|---|---|---|
| | Best train loss | Best val loss | AUROC | Avg ED (Related) | Avg ED (Unrelated) | UKBB |
| 1 | 202.2 | 323.61 | 0.89 | 15.11 | 42.1 | 0.91 |
| 2 | 277.5 | 251.51 | 0.94 | 12.91 | 44.46 | 0.924 |
| 3 | 282.2 | 314.46 | 0.901 | 14.73 | 43.2 | 0.919 |
| 4 | 324 | 317.28 | 0.903 | 16.06 | 43.43 | 0.9 |
| 5 | 201.4 | 293.68 | 0.913 | 14.68 | 43.29 | 0.909 |
| 6 | 217.7 | 277.8 | 0.92 | 13.27 | 43.55 | 0.913 |
| 7 | 248.2 | 269.99 | 0.925 | 13.26 | 42.49 | 0.918 |
| 8 | 313.2 | 344.73 | 0.891 | 14 | 40.67 | 0.896 |
| 9 | 228.9 | 268.17 | 0.923 | 13.97 | 44.07 | 0.902 |
| 10 | 239 | 297.1 | 0.904 | 14.05 | 44.57 | 0.912 |

**Supplementary Table 2.** Table showing for the 10 different cross validation folds, corresponding train and val losses, Avg Euclidean Distances of Related vs Unrelated cohorts as well as AUROC's on the respective Validation sets when the model corresponding to fold 2 is fine-tuned with the LIFE-Adult dataset.

| | age matched | | | | |
|---|---|---|---|---|---|
| LIFE-Adult Fold | TRAIN | VAL | | | |
| | Best train loss | Best val loss | AUROC | Avg ED (Related) | Avg ED (Unrelated) |
| 1 | 492 | 659.23 | 0.569 | 25.62 | 27.4 |
| 2 | 363.6 | 646.9 | 0.601 | 24.49 | 26.67 |
| 3 | 498 | 619.47 | 0.63 | 22.97 | 27.13 |
| 4 | 503.9 | 645.7 | 0.568 | 19.63 | 22.8 |
| 5 | 624.6 | 583.56 | 0.636 | 21.15 | 25.44 |
| 6 | 616.6 | 586.31 | 0.659 | 22.56 | 27.42 |
| 7 | 635.9 | 658.39 | 0.578 | 23.93 | 27.62 |
| 8 | 561.3 | 709.35 | 0.537 | 27.49 | 28.63 |
| 9 | 449 | 596.67 | 0.655 | 23.25 | 28.07 |
| 10 | 514.7 | 746.18 | 0.502 | 27.29 | 27.36 |

**Supplemental Figure 1.** (a) Dataset creation pipeline for the UKBB dataset (b) Sampling of unrelated pairs (c) Dataset creation pipeline for the LIFE-Adult dataset

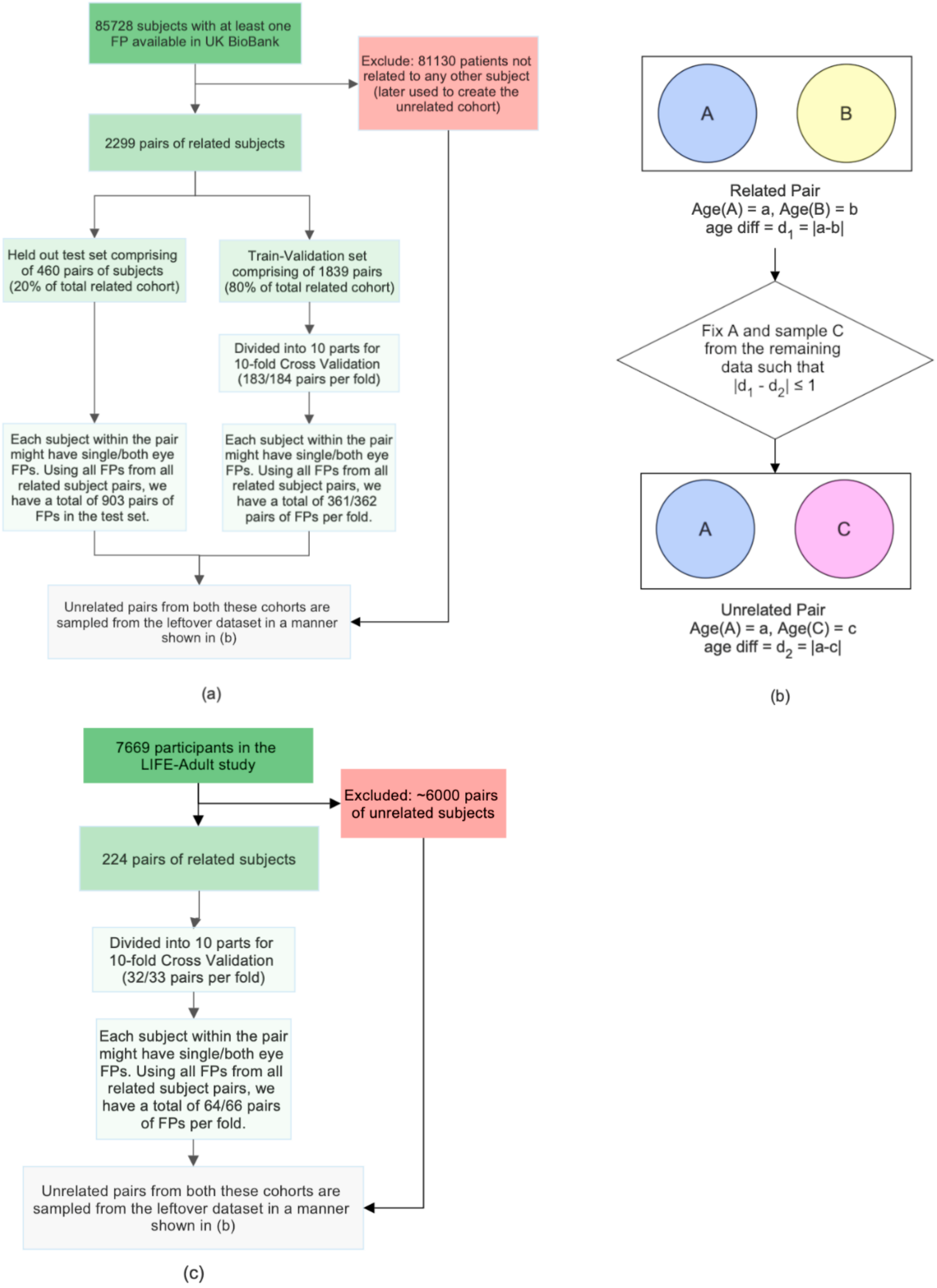

**Supplemental Figure 2.** a) Example of a UNet vessel segmentation b) ROC curve for the Siamese network trained on vessel segmented images

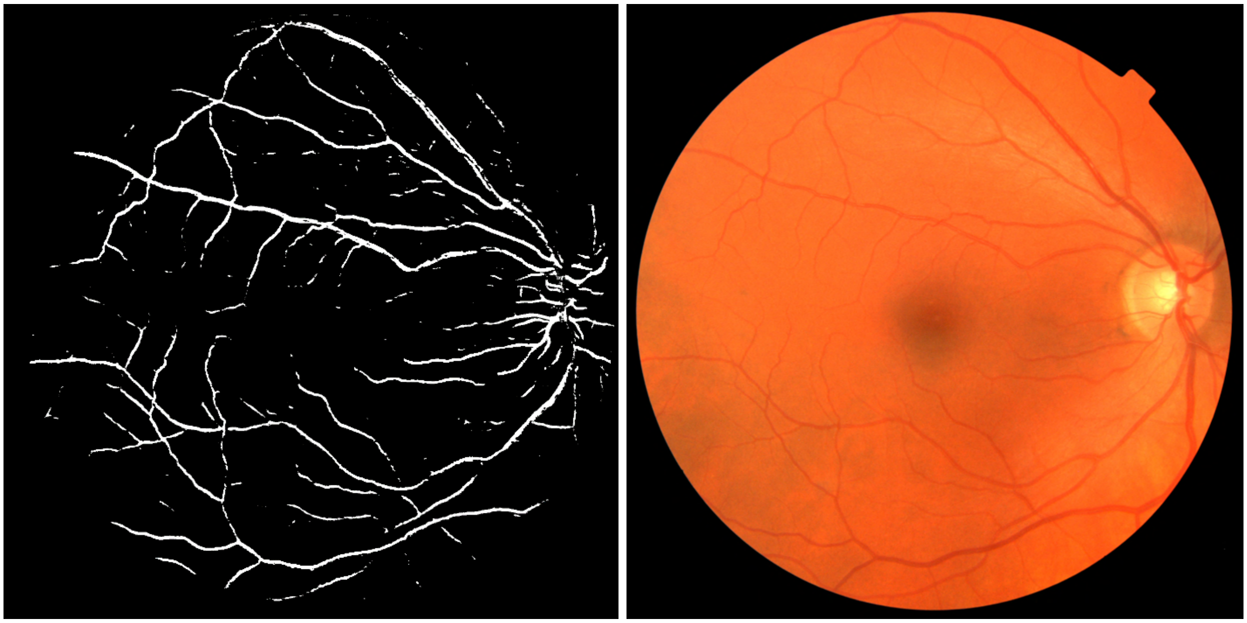

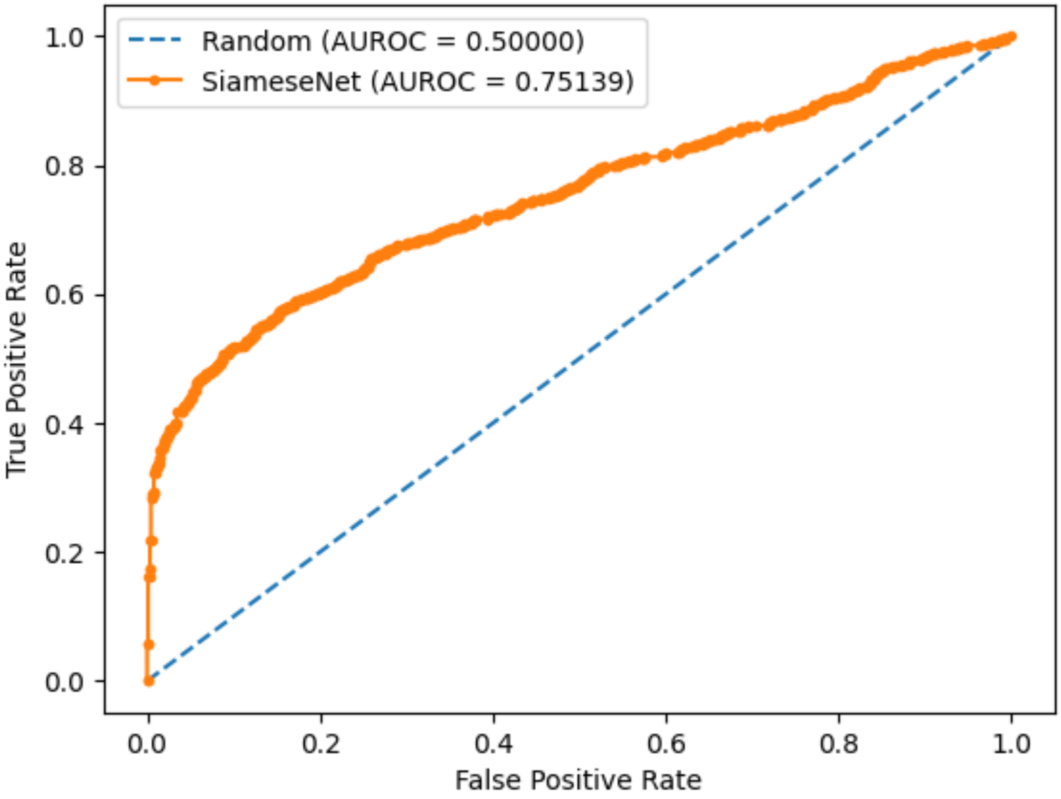

**Supplemental Figure 3.** Results after fine tuning the best-performing UKBB model (cross-validation model closest to ensemble performance) on LIFE-Adult dataset using tenfold cross validation.

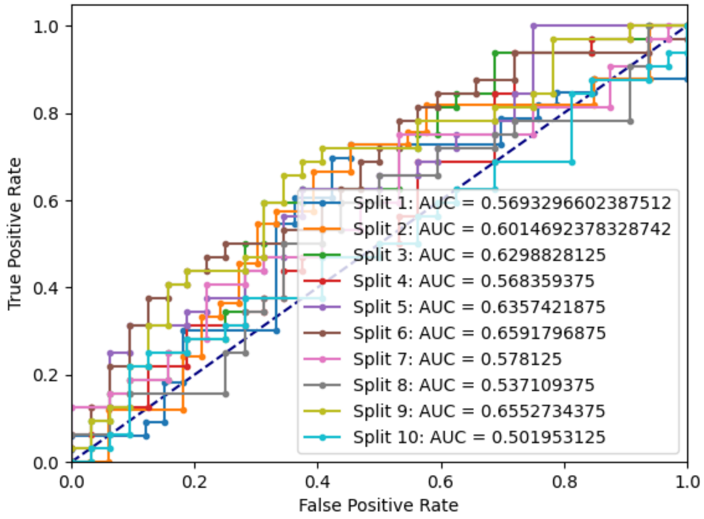
